# mRNA-1273 vaccine (Moderna): a better option than BNT162b2 (Pfizer) in kidney transplant recipients and dialysis patients?

**DOI:** 10.1101/2021.09.15.21263320

**Authors:** Veerle P. W. M. Wijtvliet, Kevin K. Ariën, Steven Abrams, Marie M. Couttenye, Fabienne Mestrez, Joachim Mariën, Benedicte Y. De Winter, Pierre Van Damme, Lissa Pipeleers, Karl Martin Wissing, Daniel Abramowicz, Kristien J. Ledeganck

## Abstract

In this study of the humoral immune response after the SARS-CoV-2 mRNA vaccination, low seroconversion rates were noted in both kidney transplant recipients and dialysis patients after first dose administration. While in DP the seroconversion rate levelled up to 92.8% after a second dose of SARS-CoV-2 mRNA vaccination, KTR stayed behind with a seroconversion rate of 62.4%. Futhermore, vaccination with the mRNA-1273 vaccine (Moderna) resulted in both higher seroconversion rates and mean log-transformed antibody titer concentrations compared to BNT162b2 (Pfizer).

## Main text

With the roll-out of COVID-19 vaccines, it has become clear that vaccinating the majority of the population worldwide will be the most important element to control and manage the ongoing COVID-19 pandemic. Several reports have shown diminished immunogenicity of SARS-CoV-2 vaccines in both kidney transplant recipients (KTR) and dialysis patients (DP), as evidenced in a recent narrative review [1]. Furthermore, two recent articles have included comparisons between the immunogenicity of the mRNA-1273 (Moderna) versus the BNT162b2 (Pfizer) vaccine in these populations [2, 3]. Boyarsky et al. studied a cohort of solid organ transplant recipients, including KTR, and reported lower seroconversion rates after receiving the Pfizer vaccine [3]. Stumpf et al. also described lower seroconversion rates in KTR and DP receiving the Pfizer vaccine [2]. However, none of these articles reported the titers of anti-SARS-CoV-2 antibodies. Therefore, we here compared the immunogenicity of the two available mRNA vaccines BNT162b2 (Pfizer-BioNTech) versus mRNA-1273 (Moderna) in both KTR and DP regarding both seroconversion rates and, among responding patients, antibody titers.

KTR and DP receiving a first dose of a SARS-CoV-2 mRNA vaccine between February and May 2021 were consecutively included. Blood was prospectively sampled at day 1 (before vaccination), 16 to 21 days after the first dose of the vaccine, and 21 to 35 days after the second vaccine dose. Samples were stored at “Biobank Antwerp”, Antwerp, Belgium; ID: BE 71030031000 [4]. Immunoglobulin G (IgG) antibodies against the receptor-binding domain (RBD) of the SARS-CoV-2 spike protein were measured using an in-house Luminex assay [5]. These measurements were performed by staff unaware of the identity of the samples. In order to investigate only patients without prior SARS-CoV-2 infection, 1) patients with a PCR-proven history of SARS-CoV-2 infection and 2) patients who tested positive for anti-RBD IgG antibodies (> 1) at the day of vaccination without a PCR-proven history of SARS-CoV-2 infection, as a surrogate of previous SARS-CoV-2 infection, were excluded. Response to first and second vaccination was defined as anti-RBD IgG positivity (signal-to-noise ratio > 1) at 16 to 21 days after dose 1, and 21 to 35 days after dose 2, respectively. The study was performed in accordance with the Declaration of Helsinki and approved by the ethical committee of the Antwerp University Hospital and University of Antwerp (no. EC 21/05/076).

Multiple logistic regression modelling was used to identify variables associated with a positive antibody response after dose 1 and dose 2. A multiple linear mixed model was used to study factors influencing the mean log-transformed anti-RBD IgG levels in responding patients. Potentially important factors were first identified using a bivariate analysis (with effect sizes expressed as crude Odds Ratios (ORs) or additive effects, respectively). Next to that, a stepwise model building exercise based on Akaike’s Information Criterion (AIC) was used to select all important explanatory variables in the final models. Depending on the selected covariates, all pairwise interaction effects were checked for significance adjusting for multiplicity. All statistical tests were two-sided. A p-value < 0.05 was considered statistically significant.

In total, 155 KTR and 182 DP were included, of whom 268 patients without prior SARS-CoV-2 infection (80%) were sampled after both vaccine dose 1 and dose 2. To evaluate the antibody response after dose 1, 138 KTR and 141 DP were included after exclusion of patients with evidence of prior SARS-CoV-2 exposure (n = 55) or loss to follow up (n = 3) (**Table 1**). Identical exclusion criteria were used to study antibody response after dose 2, resulting in the inclusion of 133 KTR and 138 DP (**Table 2**).

**Table 1:** Patient characteristics (first dose response) (univariate analysis)

|  | mRNA-1273<br>(Moderna) | BNT162b2 vaccine<br>(Pfizer-BioNTech) | p-Value |
| --- | --- | --- | --- |
| **Kidney transplant recipients (n = 138)** |  |  |  |
| Number of subjects | 44 | 94 |  |
| Male (n (%)) | 26 (59.0) | 54 (57.4) | 0.855* |
| Median age, years (IQR) | 54.0 (48.8-61.0) | 56.5 (46.0-66.8) | 0.293** |
| Diabetes mellitus (n (%)) | 3 (6.8) | 11 (11.7) | 0.548* |
| Immunosuppressive therapy (n (%)) |  |  |  |
| CNI | 6 (14.6) | 23 (24.4) | 0.146* |
| CNI + MMF/AZA/mTOR inhibitor | 34 (77.3) | 58 (61.7) | 0.071* |
| Positive antibody response (n (%)) | 11 (25.0) | 8 (8.5) | **0.009\*** |
| Anti-RBD IgG antibody level†<br>(median (IQR)) | 3.13 (1.93-7.83) | 5.03 (3.03-6.71) | 0.386** |
| Anti-RBD IgG antibody level†<br>(geometric mean (GSD)) | 3.77 (2.36) | 4.80 (2.06) | 0.529*** |
| **Dialysis patients (n = 141)** |  |  |  |
| Number of subjects | 61 | 80 |  |
| Male (n (%)) | 39 (63.9) | 53 (66.3) | 0.775* |
| Median age, years (IQR) | 70 (60.0-78.0) | 70 (62.0-75.0) | 0.975** |
| Haemodialysis (n (%)) | 40 (65.6) | 63 (78.8) | 0.081* |
| Diabetes mellitus (n (%)) | 29 (47.5) | 28 (35.0) | 0.133* |
| Positive antibody response (n (%)) | 41 (67.2) | 21 (26.3) | **< 0.001\*** |
| Anti-RBD IgG antibody level†<br>(median (IQR)) | 6.43 (3.28-10.54) | 3.03 (1.96-5.03) | **0.011\*\*** |
| Anti-RBD IgG antibody level†<br>(geometric mean (GSD)) | 5.27 (2.26) | 3.17 (1.91) | **0.016\*\*\*** |
| **Total (n = 279)** |  |  |  |
| Number of subjects | 105 | 174 |  |
| Male (n (%)) | 65 (61.9) | 107 (61.4) | 0.946* |
| Median age, years (IQR) | 61.0 (50.0-72.0) | 63.0 (51.5-71.0) | 0.881** |
| Diabetes mellitus (n (%)) | 32 (30.4) | 39 (22.4) | 0.134* |
| Positive antibody response (n (%)) | 52 (49.5) | 29 (16.7) | **< 0.001\*** |
| Anti-RBD IgG antibody level†<br>(median (IQR)) | 5.70 (2.37-10.52) | 3.34 (2.05-5.96) | 0.054** |
| Anti-RBD IgG antibody level†<br>(geometric mean (GSD)) | 4.91 (2.29) | 3.55 (1.98) | 0.077*** |

† In responding patients
\* Based on Pearson's Chi-square test (or Fisher exact test in case of small cell counts)
\*\* Based on Mann-Whitney U test
\*\*\* Based on an unpaired Student's t-test comparing two mean log-GMTs
*Abbreviations:* AZA = azathioprine; CS = corticosteroids; CNI = calcineurin inhibitors; GSD = geometric standard deviation; IgG = immunoglobulins G; IQR = interquartile range; MMF = mycophenolate mofetil; mTOR = mechanistic target of rapamycin; n = number; RBD = receptor binding domain; SD = standard deviation

**Table 2:** Patient characteristics (second dose response) (univariate analysis)

|  | mRNA-1273<br>(Moderna) | BNT162b2 vaccine<br>(Pfizer-BioNTech) | p-Value |
| --- | --- | --- | --- |
| **Kidney transplant recipients (n = 133)** |  |  |  |
| Number of subjects | 42 | 91 |  |
| Male (n (%)) | 25 (59.5) | 52 (57.1) | 0.796* |
| Median age, years (IQR) | 55.5 (50.0-61.0) | 57.0 (46.5-66.5) | 0.365** |
| Diabetes mellitus (n (%)) | 3 (7.1) | 10 (11.0) | 0.754* |
| Immunosuppressive therapy (n (%)) |  |  |  |
| CNI | 6 (14.3) | 22 (24.2) | 0.193* |
| CNI + MMF/AZA/mTOR inhibitor | 33 (78.6) | 56 (61.5) | 0.052* |
| Positive antibody response (n (%)) | 32 (76.2) | 51 (56.0) | **0.026\*** |
| Anti-RBD IgG antibody level†<br>(median (IQR)) | 12.39 (3.51-16.18) | 5.62 (2.13-14.25) | 0.159** |
| Anti-RBD IgG antibody level†<br>(geometric mean (GSD)) | 7.27 (2.63) | 5.37 (2.73) | NA*** |
| **Dialysis patients (n = 138)** |  |  |  |
| Number of subjects | 58 | 80 |  |
| Male (n (%)) | 35 (60.3) | 53 (66.3) | 0.476* |
| Median age, years (IQR) | 69.5 (60.0-77.8) | 70.0 (61.8-75.0) | 0.941** |
| Haemodialysis (n (%)) | 37 (63.8) | 63 (78.8) | 0.052* |
| Diabetes mellitus (n (%)) | 25 (43.1) | 29 (36.3) | 0.416* |
| Positive antibody response (n (%)) | 58 (100) | 70 (87.5) | **0.005\*** |
| Anti-RBD IgG antibody level†<br>(median (IQR)) | 16.16 (15.69-16.41) | 16.58 (10.22-17.36) | 0.194** |
| Anti-RBD IgG antibody level†<br>(geometric mean (GSD)) | 15.22 (1.23) | 11.86 (1.98) | NA*** |
| **Total (n = 271)** |  |  |  |
| Number of subjects | 100 | 171 |  |
| Male (n (%)) | 60 (60.0) | 105 (61.4) | 0.819* |
| Median age, years (IQR) | 61.0 (50.8-72.0) | 63.0 (53.0-71.0) | 0.831** |
| Diabetes mellitus (n (%)) | 28 (28.0) | 39 (22.8) | 0.339* |
| Positive antibody response (n (%)) | 90 (90.0) | 121 (70.8) | **< 0.001\*** |
| Anti-RBD IgG antibody level†<br>(median (IQR)) | 16.02 (13.42-16.39) | 14.08 (4.89-17.14) | 0.449** |
| Anti-RBD IgG antibody level†<br>(geometric mean (GSD)) | 11.70 (2.00) | 8.49 (2.51) | NA*** |

† In all responding patients after second vaccine dose
\* Based on Pearson's Chi-square test (or Fisher exact test)
\*\* Based on Mann-Whitney U test
\*\*\* No p-values were determined for geometric mean anti-RBD IgG antibody levels, given deviations from normality
*Abbreviations:* AZA = azathioprine ; CS = corticosteroids ; CNI = calcineurin inhibitors ; GSD = geometric standard deviation; IgG = immunoglobulins G ; IQR = interquartile range ; MMF = mycophenolate mofetil; mTOR = mechanistic target of rapamycin; n = number ; NA = not applicable; RBD = receptor binding domain ; SD = standard deviation

When zooming in on patients included to study antibody response after dose 1 (n = 279), the median age was 62 years (interquartile range (IQR), 51-71 years) and 62% were men. Sixty-two percent received the BNT162b2 vaccine (Pfizer-BioNTech), 38% received the mRNA-1273 vaccine (Moderna). At a median of 21 days (IQR, 20-23 days) anti-RBD IgG antibodies were detected in 81 out of 279 participants (29%, Clopper-Pearson 95% CI, 24-35%). Both haemodialysis patients (adjusted OR, 6.403 (95% CI, 2.970-13.804); p < 0.001) and peritoneal dialysis patients (adjusted OR, 10.379 (95% CI, 4.143-26.000; p < 0.001) were more likely to develop antibody responses compared to KTR (**Table 3A**). Furthermore, the odds of having an antibody response decreased with a factor 0.972 (95% CI, 0.950-0.994) with a one-year increase in age. Patients receiving the mRNA-1273 vaccine (Moderna) showed significantly different seroconversion rates 21 days after dose 1 compared to patients receiving the BNT162b2 vaccine (Pfizer-BioNTech) (49.5% versus 16.7%; adjusted OR, 4.941 (95% CI, 2.698-9.047); p < 0.001) (**Table 1 and Table 3A**). All pairwise interaction terms included in both models were found to be non-significant at a 5% significance level. Performing a similar analysis regarding antibody responses after second vaccine dose, anti-RBD IgG antibodies were detected in 211 out of 271 participants (78%, Clopper-Pearson 95% CI, 72-83%) at a median of 28 days (IQR, 25-31 days) after second vaccine administration. Again, both haemodialysis patients (adjusted OR, 6.181 (95% CI, 2.828-13.511); p < 0.001) and peritoneal dialysis patients (adjusted OR, 18.870 (95% CI, 2.483-143.380; p = 0.005) were more likely to develop antibody responses compared to KTR (**Table 3A**). Patients receiving the mRNA-1273 vaccine (Moderna) showed significantly different seroconversion rates compared to those receiving the BNT162b2 vaccine (Pfizer-BioNTech) (90.0% versus 70.8%; adjusted OR, 3.428 (95% CI, 1.587-7.406); p < 0.005) (**Table 2 and Table 3A**). After selecting patients with an antibody response after first (n = 81) or second (n = 211) vaccine dose, antibody levels appeared to be associated with type of patient, age and vaccine type. Mean log-transformed antibody titer concentrations were 0.289 units higher for Moderna vs. Pfizer vaccination, irrespective of the dose (first or second) considered (p = 0.005 in multiple linear mixed model; **Table 3B**).

**Table 3.**
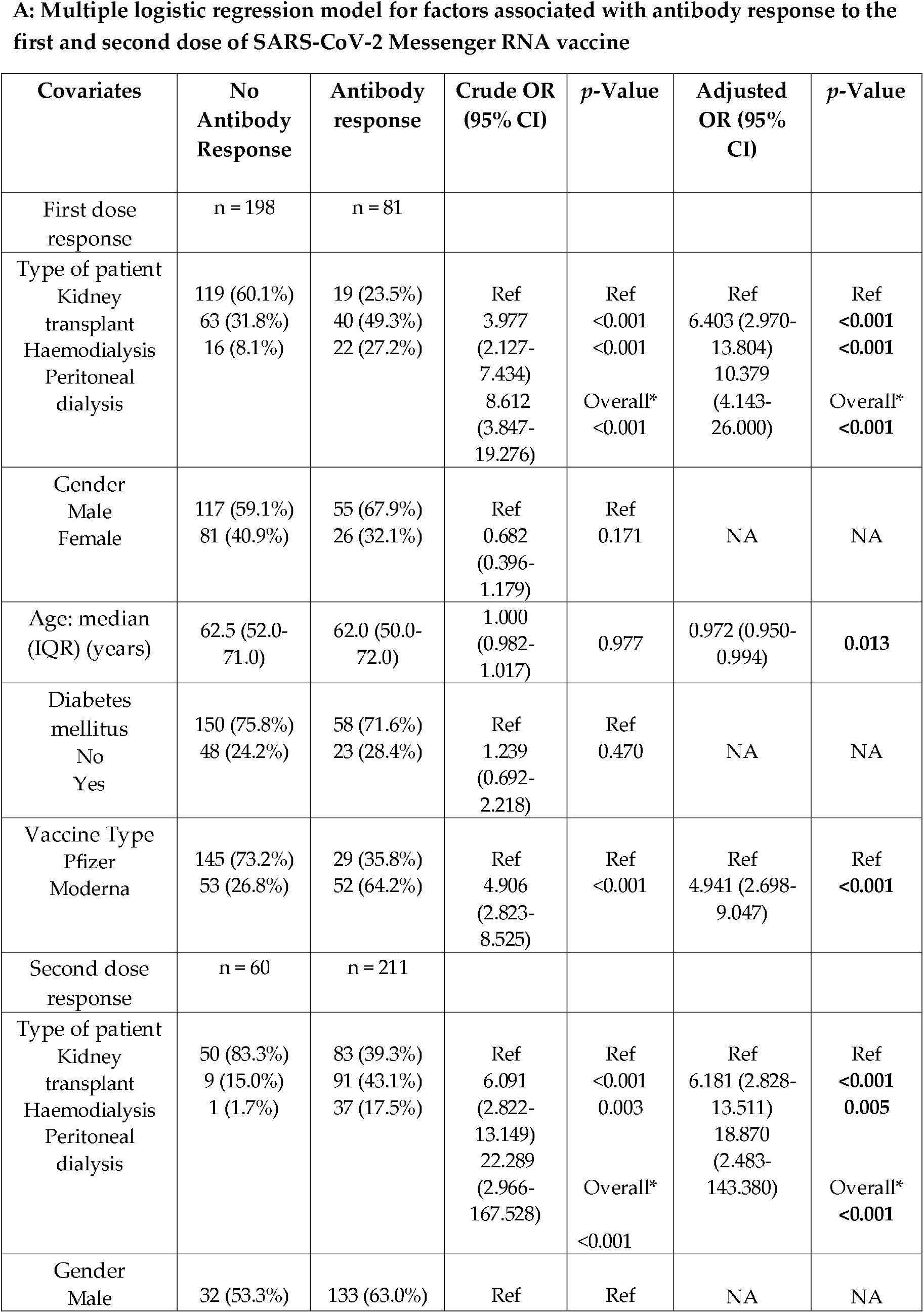
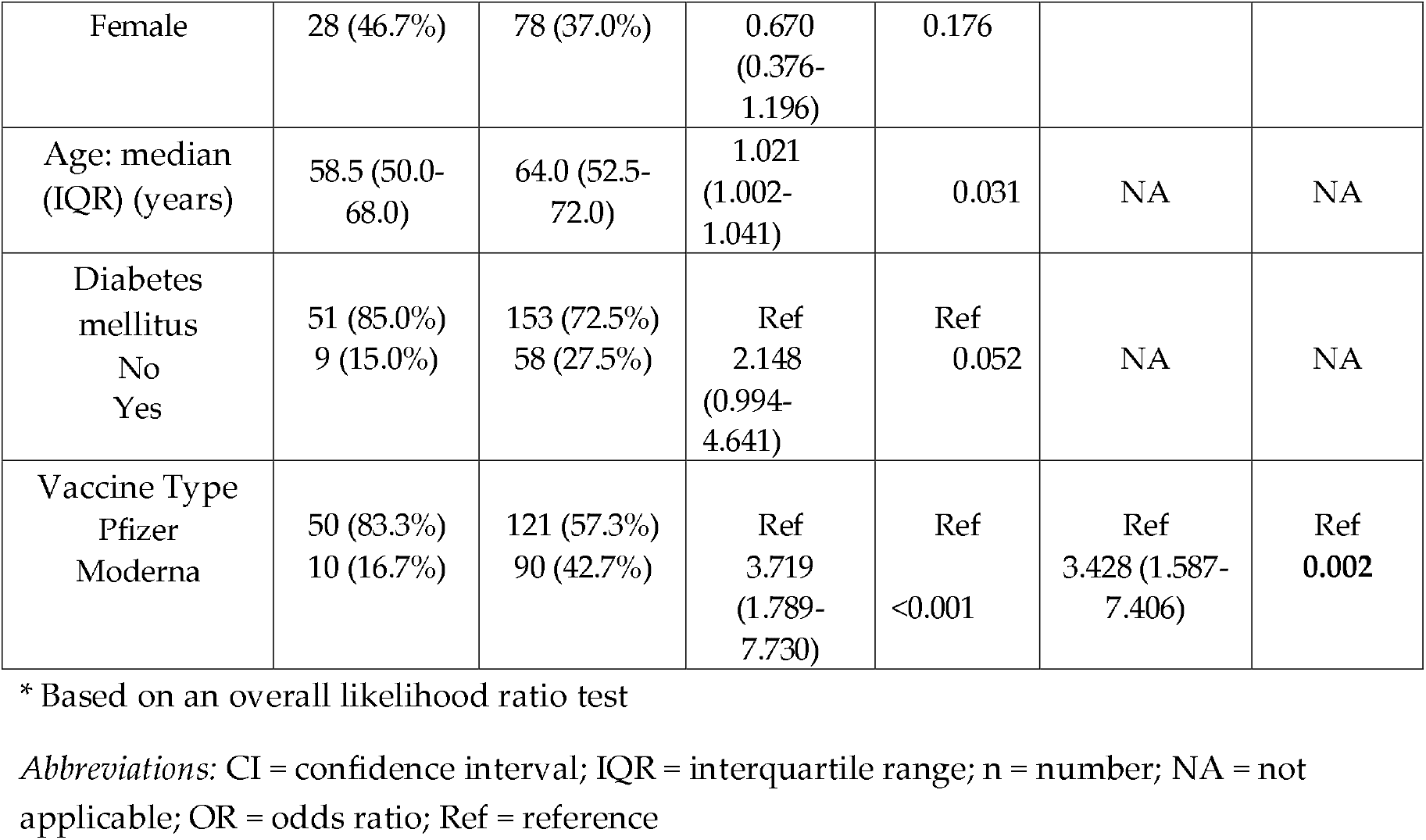
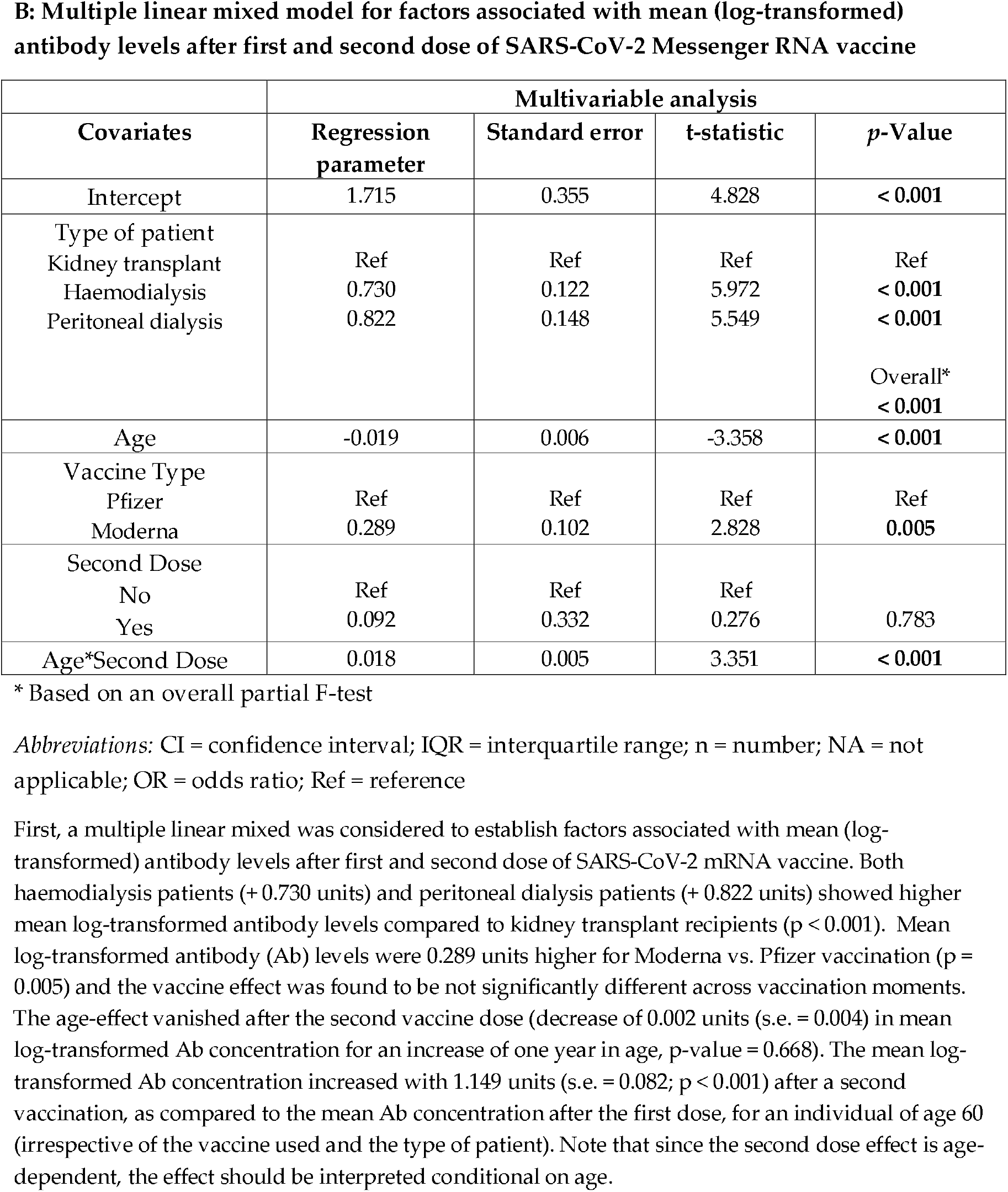

In this study of the humoral immune response after SARS-CoV-2 mRNA vaccination, low seroconversion rates were noted in both KTR and DP after first dose administration. While in DP the seroconversion rate levelled up to 92.8% after a second dose of SARS-CoV-2 mRNA vaccination, KTR stayed behind with a seroconversion rate of 62.4%. Furthermore, vaccination with the mRNA-1273 vaccine (Moderna) resulted in both higher seroconversion rates and mean log-transformed antibody titer concentrations.

Several limitations of our study, must be acknowledged. First, although an interesting difference in serological response was observed following administration of mRNA-1273 vs. BNT162b2, no further investigation was performed on potential differences in T-cell responses. It is well described in literature that patients without antibody responses can mount T-cell responses, which could provide some protection against SARS-CoV-2 infection in seronegative patients [6]. Second, no information concerning possible side effects of both SARS-CoV-2 mRNA vaccines, was collected.

While writing this discussion, three very recently articles were published, which support our above-mentioned findings [7-9]. First, seroconversion rates in adults with hematologic malignant disease, whom in majority received B-cell-depleting monoclonal antibodies, were higher in patients vaccinated with mRNA-1273 (Moderna) compared to BNT162b2 (Pfizer) (50% vs. 33%, p = 0.013) [7]. Second, a study of individuals in the multi-state Mayo Clinic Health System revealed that the mRNA-1273 vaccine was associated with a lower incidence of SARS-CoV-2 infection (p = 0.0034) as well as with a significant two-fold risk reduction against breakthrough infection compared to BNT162b2 [8]. Third, Kaiser et al. compared antibody titers between both mRNA vaccines after full vaccination in haemodialysis patients [9]. They reported 2.39-fold (P < 0.005) higher anti-S-antibody titers in patients vaccinated with mRNA-1273.

Whether observed differences between these two mRNA-vaccines relate to a different mRNA sequence, a difference in dosage (100 μg mRNA in Moderna vs. 30 μg mRNA in Pfizer) or a difference in vaccine composition, remains to be investigated. Furthermore, future studies are needed to assess whether our observed differences are associated with a better clinical protection. Meanwhile, mRNA-1273 (Moderna) might be the preferred vaccine in kidney transplant and dialysis patients.

## Data Availability

Requests for original and additional data should be directed to the corresponding author.

## Acknowledgements

The authors would like to thank study nurses E. Meersman, S. Verhofstede, N. Marmitte and T. Bogaerts for their excellent and efficient help. We also thank E. Snelders, S. Van Hees and J. Michiels for their logistical support, the lab technicians of the Lab of Experimental Medicine and Pediatrics for the sample processing and P. Moons from Biobank Antwerp for his technical support. Special thanks to all the kidney transplant recipients and dialysis patients who made this study possible.

## Data availability statement

Requests for original and additional data should be directed to the corresponding author.

